# Temperature and Humidity Do Not Influence Global COVID-19 Incidence as Inferred from Causal Models

**DOI:** 10.1101/2020.06.29.20142307

**Authors:** Raghav Awasthi, Aditya Nagori, Pradeep Singh, Ridam Pal, Vineet Joshi, Tavpritesh Sethi

**Affiliations:** Indraprastha Institute of Information Technology Delhi, India; All India Institute of Medical Sciences, New Delhi, India; CSIR-Institute of Genomics and Integrative Biology, Delhi, India

## Abstract

The relationship between meteorological factors such as temperature and humidity with COVID-19 incidence is still unclear after 6 months of the beginning of the pandemic. Some literature confirms the association of temperature with disease transmission while some oppose the same. This work intends to determine whether there is a causal association between temperature, humidity and Covid-19 cases. Three different causal models were used to capture stochastic, chaotic and symbolic natured time-series data and to provide a robust & unbiased analysis by constructing networks of causal relationships between the variables. Granger-Causality method, Transfer Entropy method & Convergent Cross-Mapping (CCM) was done on data from regions with different temperatures and cases greater than 50,000 as of 13^th^ May 2020. From the Granger-Causality test we found that in only Canada, the United Kingdom, temperature and daily new infections are causally linked. The same results were obtained from Convergent Cross Mapping for India. Again using Granger-Causality test, we found that in Russia only, relative humidity is causally linked to daily new cases. Thus, a Generalized Additive Model with a smoothing spline function was fitted for these countries to understand the directionality. Using the combined results of the said models, we were able to conclude that there is no evidence of a causal association between temperature, humidity and Covid-19 cases.

## Introduction

Ever since the outbreak of coronavirus at the global stage and WHO declaring it as pandemic on 11th February 2020, multiple speculations started surfacing for controlling this outbreak. Prediction for active cases turned out to be the most highlighted research, as flattening the curve was a primary goal everyone was aiming for. Various Mathematical and Artificial Intelligence based models [6,7,8,9] were used for this task to provide government, health advisories and policy makers substantial information that would help them make decisions. No study has been done so far that incorporates the effect of environmental factors in these models. These factors might not only help us improve the general fit of the predictive model but also provide us with a deeper insight into ways of curbing the spread. Despite the above, there is no conclusive evidence in the past literature which states whether temperature or humidity play a role in affecting the infection rate. The research community had diverse opinions on these two factors. In this study we tried to address this issue and examined the causal association of temperature and humidity with the spread of COVID-19 pandemic. Jingui et. al stated that coronavirus is positively affected with temperature[1]. The Generalized Additive Model (GAM) was used to show association between temperature and rate of infection in this paper [2]. Another significant study was done to portray the effects of temperature and humidity in death rates for COVID patients in Wuhan, China stating that humidity was negatively correlated with COVID [2]. Even in this study the GAM model was used to depict the association of humidity with the infection rate. Ye Yao et. al stated in his research that there is no significant association of COVID-19 with temperature and UV radiations in Chinese cities; exploring the association through various regression methods[3]. Investigations done by Jingyuan et. al suggested that high temperature and high humidity reduces the transmission of COVID-19 based on cross sectional regression analysis.[4]. Extending ideas from the above, our study investigates various causal effects of temperature and humidity on active COVID-19 cases for selected countries around the world. Causal Inference can state how one factor can transmit its effect on other factors, and also make connection of an effect with its cause. We have used three different causality tests for finding these critical insights and state conclusively whether temperature and humidity have an effect in the rate of infections or not.

## Methods

### Dataset

The environmental data was downloaded from https://power.larc.nasa.gov/data-access-viewer/ and the Covid-19 data was taken from https://github.com/CSSEGISandData/COVID-19. Using the same, we extracted the relative humidity and average temperature in 2M and country wise count of daily new cases for our analysis.

### Location Identification for the Analysis

From every 7 regions defined by WHO we selected only those countries which have more than 50,000 cases till 13th May and show considerable variations in their average temperature.

### Analysis

In order to get robust and unbiased results we performed multiple causal tests on our data. Granger causality, Transfer Entropy (suitable for stochastic systems) and Convergent Cross-Mapping (suitable for Dynamical systems) were used for calculating the causal relationships. Further, for countries where we had a significant amount of causal relationship we used Generative Additive Model for calculating the directionality of the same.

### Granger-Causality

The first test of causal association was performed using the Autoregressive based Granger-Causality method. The Granger-Causality method is used to find if one time-series can be the cause for another or vice-versa. in-order to optimize lag p (i.e. past p values) of the time series we fit an AR (p) (where p varies from 1 to 15) model for every time series and then identify lag with minimum BIC (Bayesian information criteria) score) [6]. The Granger-Causality method is mainly useful for analyzing numerical time series data produced by processes having patterns which are analyzed statistically. But a key limitation of this technique is that it is restricted to stationary time-series data. We performed our analysis with R Package lmtest[13].

**Table1:** Country-wise P-values obtained from Granger-causality Test table shows value upto 3 decimal places.

| Country | P-value Temperature Causes New Cases | P-value Humidity Causes New Cases |
| --- | --- | --- |
| Belgium | 0.127 | 0.007 |
| Brazil | 0.011 | 0.151 |
| Canada | 0.002 | 0.014 |
| China | 0.007 | 0.886 |
| France | 0.505 | 0.049 |
| Germany | 0.665 | 0.249 |
| India | 0.592 | 0.204 |
| Iran | 0.092 | 0.571 |
| Italy | 1 | 0.461 |
| Peru | 0.026 | 0.323 |
| Russia | 0.074 | 0 |
| Spain | 0.784 | 0.627 |
| Turkey | 0.492 | 0.132 |
| United Kingdom | 0.003 | 0.008 |
| US | 0.019 | 0.288 |

### Transfer-Entropy

Transfer-Entropy is used to measure the amount of information that gets transferred from one variable to another. Transfer-Entropy helps us formalize the amount of uncertainty reduced in the future values of a time series by knowing the past values of another. In this analysis, we calculated the Shannon entropy [17] for this task by assuming the time-series as a sequence of discrete symbols and then running the model for 300 bootstraps. Significant causality between variables based on p values (<0.05) was assumed and then the lag optimization was performed same as mentioned above. R software RtransferEntropy was used for the analysis [10].

**Table2:** Country-wise P-values obtained from Transfer Entropy table shows value up to 3 decimal places.

| **Country** | **P-value Temperature Causes New Cases** | **P-value Humidity Causes New Cases** |
| --- | --- | --- |
| Belgium | 0.024 | 0.04 |
| Brazil | 0.008 | 0.06 |
| Canada | 0.067 | 0.105 |
| China | 0.061 | 0.055 |
| France | 0.022 | 0.052 |
| Germany | 0.058 | 0.046 |
| India | 0.121 | 0.093 |
| Iran | 0.035 | 0.029 |
| Italy | 0.099 | 0.07 |
| Peru | 0.073 | 0.068 |
| Russia | 0.151 | 0.156 |
| Spain | 0.191 | 0.094 |
| Turkey | 0.103 | 0.046 |
| United Kingdom | 0.021 | 0.053 |
| US | 0.065 | 0.039 |

### Convergent Cross-Mapping

The Transfer-Entropy and the Granger-Causality techniques outlined above are suitable only for the systems having probability distribution. But one may also encounter numerical data that may have originated from a Deterministic Dynamical System. Coupled differential equations can be used to model such systems. However, there are many cases where only a few variables which are relevant to the system are available for the researchers. To tackle this, Taken’s Theorem [11] known as Phase-State Reconstruction was used, which states that the properties of dynamical system’s attractor can be recovered using a single one of its variables. Using this theorem, Sugihara et al. (2012) developed the new mapping technique named Convergent-Cross Mapping (CCM) [12], this enabled the recovery of direction of causality between any two time sequences that are generated by the same deterministic dynamical system. To identify causal association, we embedded our time series in 3 dimensions series. And from the embedded time series we created a library and randomly sampled 100 samples from each library. Pearson correlation is used to score the cross-mapping performance, where the correlation was assumed to be greater than 0.80 for significant causal association. rEDM R package used to perform this analysis [14].

### Generalized Additive Model (GAM) for causally related time-series

We finally plotted and estimated the effect size of temperature and humidity on the countries where the variables were found to be causally related. A smoothing spline function is fitted between lagged weather conditions and log scale daily new cases. The spline curves thus obtained were plotted. Further an adjusted model with previous day’s counts data and temperature and relative humidity was built for only countries that have causal relationships as calculated from the above methods.

## Results

First using BIC score we calculated the minimum lag of Temperature, Humidity and New Cases time series (Supplementary figure 1) we obtained that in the lag range of 1-15 time series have almost similar BIC Score, so we picked the lag value 1 for our analysis. Then to check stationarity in time series we performed the Augmented Dickey Fuller test (ADF Test) and we found Temperature, Humidity and new cases time series as stationary in every country (Supplementary Table 1).

### Granger Causality

We performed an F-test to verify the Null Hypothesis i.e. Temperature and Humidity Granger Causes the new infected cases of covid-19 infected cases. For Canada and the United Kingdom temperature was found Granger causal with daily new cases with P-values 0.002 (<0.005) and 0.003(<0.005) respectively and humidity was found Granger causal with daily new cases in Russia with P-values (<0.005). For other Countries we have not found a significant P-value to explain that temperature and humidity Granger Causes daily infection spread.

### Transfer entropy

We interpreted our results using Shannon’s Entropy to depict how temperature and humidity are causally associated with daily count of cases. We found that in each country the value of p is greater than 0.05 which ensures that temperature does not have any causal effect on infection. Similarly in case of humidity the same trend can be seen.

### Convergent Cross Mapping

We embed the time series in 3 dimensions and randomly sampled 100 samples from each embedded time series and calculated the cross mapping. To interpret the results, we aggregated the cross map performance at different library sizes (varies from 10-80, with the difference of 10), which computes a mean value at each unique. Since average cross map skill found to be less than 0, meaning prediction skills are next to nothing. (Observations should not be anticorrelated with predictions), so we set negative values to 0 when plotting. Among all countries only for India we found the causal association of temperature and daily new cases and humidity wa found not causally associated for any states..

### Summary of Models

Finally below table presents the summary of results obtained from all the three models. Transfer Entropy method shows no causal relationship of cases with both temperature and humidity. Granger causality method found that temperature causes new cases for Canada and the United Kingdom whereas humidity causes new cases for Russia. CCM saw a causal relationship only for India.

**Table3:**
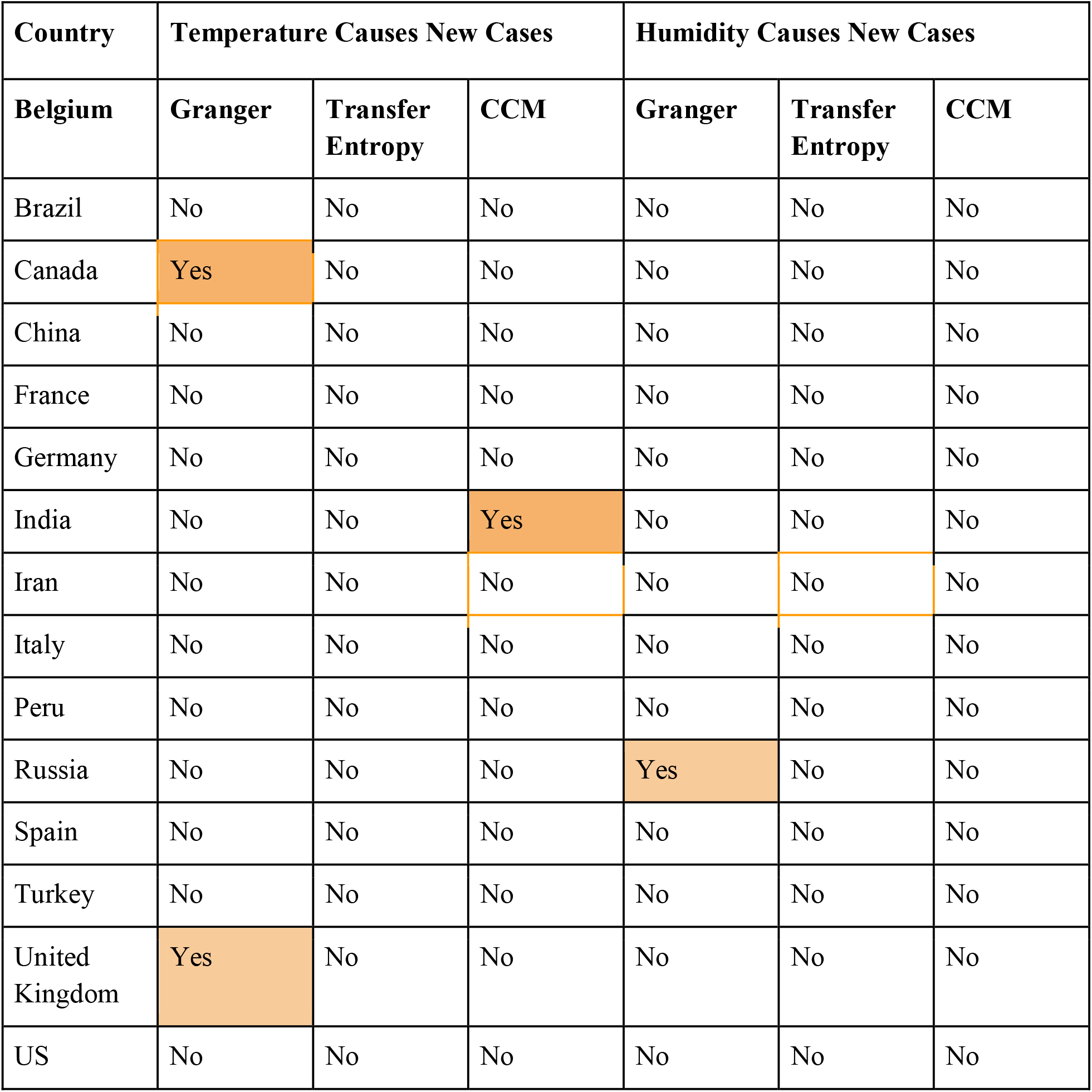
Summary Table of all models.

### Generalized Additive model results for world

From the causality model, it’s found that three countries Canada, India and the United Kingdom have causal associations between average temperature and covid19 new cases. And Russia has the causal associations between relative humidity and covid19 new cases. To reassert these relationships we looked at generalized additive models based spline fitting. For Canada, India and the UK, new cases were found to be increasing with rising temperature. For Russia, Initially new cases were found to be decreasing with increasing relative humidity but later the trend reversed i.e. increasing relative humidity resulted into increasing new cases.

## Discussion

Infection of coronavirus is exponentially expanding all around the world, which is a threat for human life. India being a populous and dense country faces a higher risk of spread. However policy interventions such as lockdown at the very beginning kept the spread linear in India. While medical facilities, policies interventions have been implemented, it has been a general question since the very beginning that can temperature and humidity be of any help in reducing the spread. This has been observed in other pandemics such as Influenza spread in 2009[15] that, with increase in the temperature and humidity the spread reduced. Also for SARS-COV1 spread it has been reported that dry and cold air can lead to the spread. For Sars-cov2 as well, many studies have happened which assert the association between weather conditions and the spread of the virus. However, in our knowledge, none of these efforts involve study of the causal effect of weather on the spread, mere associations can mislead the policy makers and can cost millions of lives. So it becomes very important for us now to estimate if there exist a causal relationship of weather conditions on infection spread and at what rate.

In this paper, we performed causal study between weather conditions with temperature and relative humidity with daily count of new infection. To involve large variability of the weather conditions, we have involved different geographical and political locations of the countries as shown in figure1. We employed a comprehensive statistical analysis to estimate the causal relationship between the time-series. It turned out the majority of the countries does not hold the significant causal relationships as computed using Granger Causality, transfer entropy and cross-convergent mapping, however a few were found to have causal relationships between weather conditions and new cases of covid19. Analysis was carried out separately for each of these in order to account for latent variability hidden within geographical and political diversity. Finally, we fitted a smoothing spline function using generalized additive models using Poisson regression to see direction of relationships. In general, most of the countries were found to have no-causal relationships. We looked at the different Geographies according to the division done by WHO [19] for different counties, we found that Canada, India and the United Kingdom were having an increased number of cases with increase in the temperature. The directionality found here is opposite to what is reported in literature for Influenza. Therefore, it would not be recommendable to take policy decisions on the basis of temperature and relative humidity as of now.

**Figure1:**
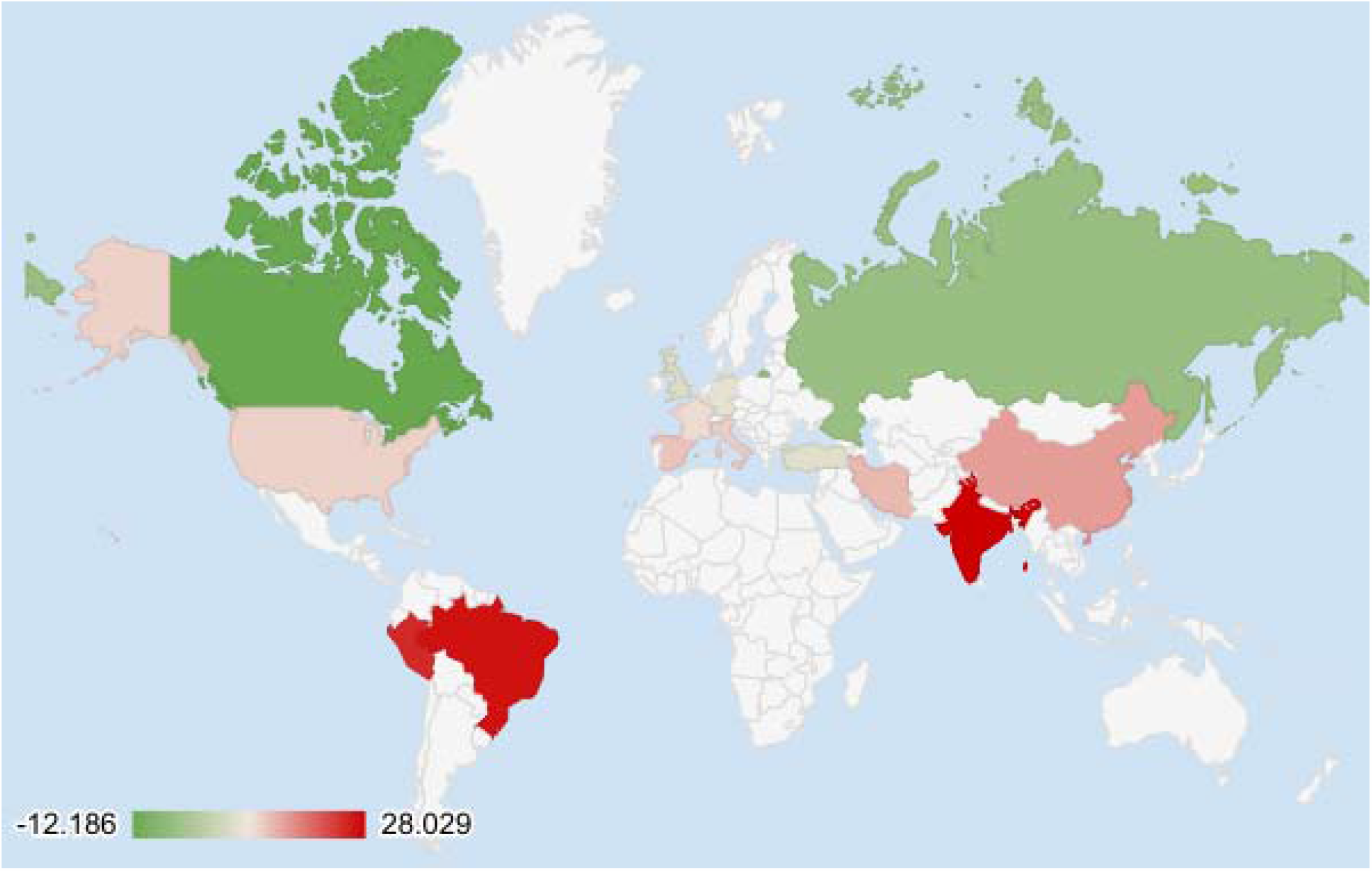
Countries with Green or Blue color are selected for the analysis and their average temperature from 30th January to 18th May.

**Figure2:**
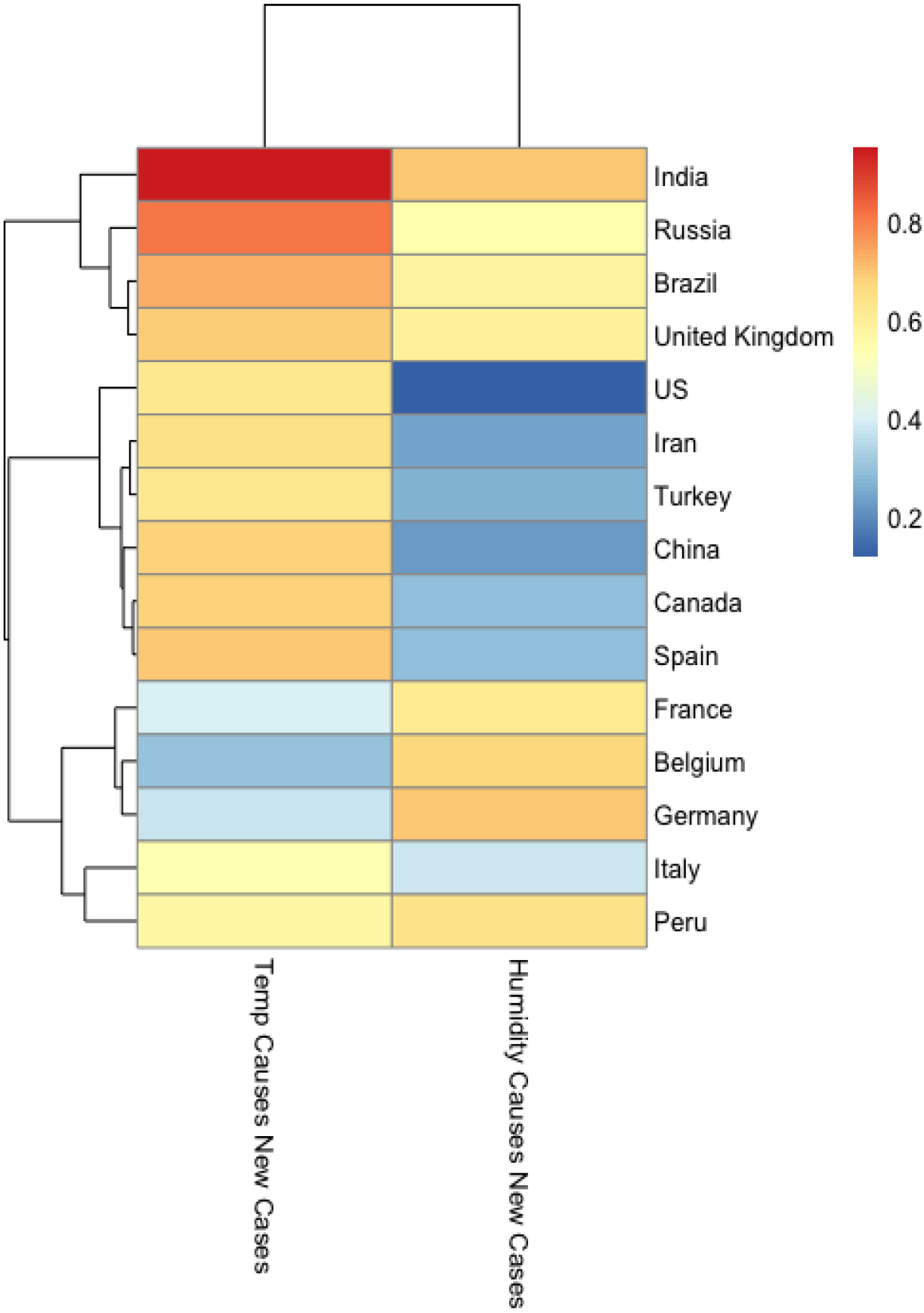
Heatmap represents Pearson correlation between values generated by cross mapping of Temperature and Humidity state space to Infection State space and original value of infection.

**Figure4:**
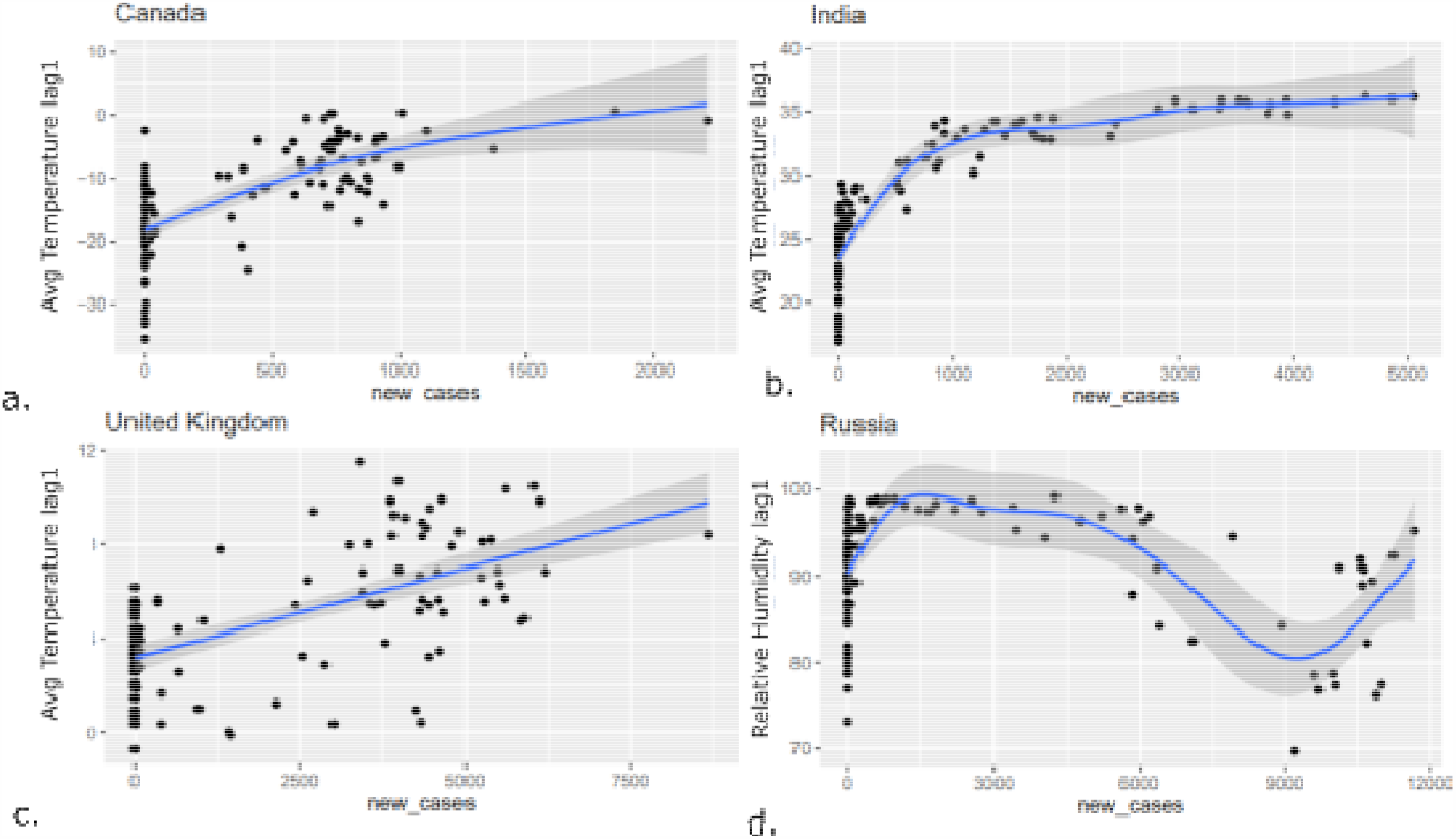
The smooth function fitted to temperature vs. new cases at lag 1 for causally related time-series at (a.) Canada, (b.) India, (c.) United Kingdom, (d.) Russia.

Our study has few limitations, it’s difficult to compute the exposure when it comes to weather conditions as many people are inside their houses due to lockdown and would avoid outdoor exposure. We also think that we have a very short period of observational data and with more data, findings may fluctuate to other sides. Overall we tried to put forth an exhaustive search for causal relationships between weather conditions and new cases of covid19 based on the available literature on causal time-series analysis and these findings asserts that we didn’t have causal association between temperature and humidity with new cases of covid19.

## Data Availability

All data used in this manuscript are publicly available.

https://power.larc.nasa.gov/data-access-viewer/

https://github.com/CSSEGISandData/COVID-19.

## Acknowledgements

This work was partially supported by the Wellcome Trust/DBT India Alliance Fellowship IA/CPHE/14/1/501504 awarded to Dr. Tavpritesh Sethi and the Center for Artificial Intelligence at IIIT-Delhi. Tavpritesh Sethi and Pradeep Singh also acknowledge the support received from the Department of Science and Technology vide project DST/INT/ISR/P-21/2017. Authors also acknowledge Prof. Rakesh Lodha from All India Institute of Medical Sciences, New Delhi, for his valuable inputs.We also thank CSIR for supporting Mr. Aditya Nagori and UGC to support Mr. Raghav Awasthi with PhD fellowships.

## Supplementary Material

### 1.) BIC calculation for lag-wise relationship estimation

A lower BIC value represents higher association than the large BIC values.

**Supplementary Figure 1:**
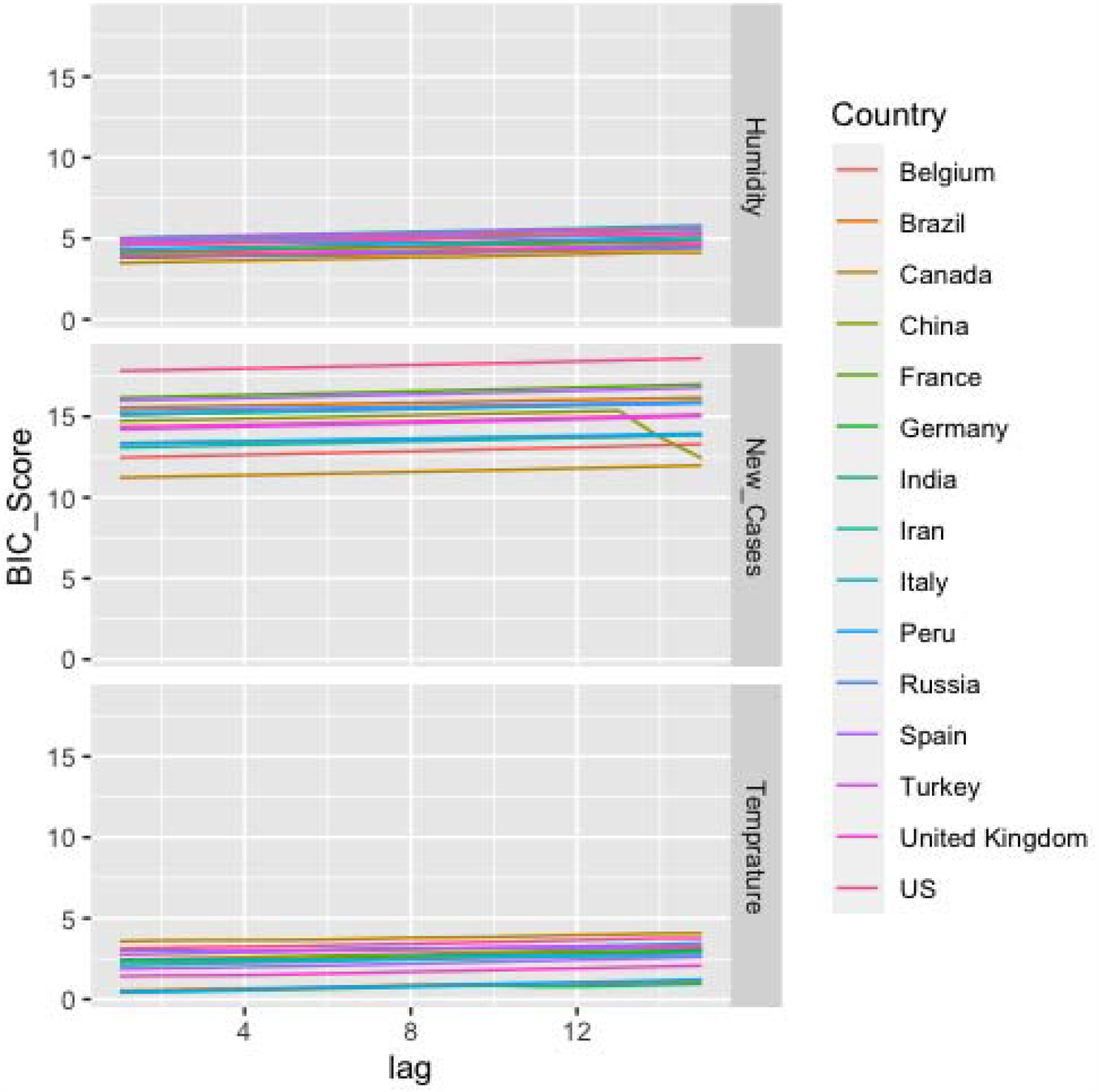
lag optimization for Temperature, Humidity, New cases time series we calculated BIC score of AR (p) model at lag 1-15.

### 2.) Augmented Dickey–Fuller Test

**Supplementary Table 1:**
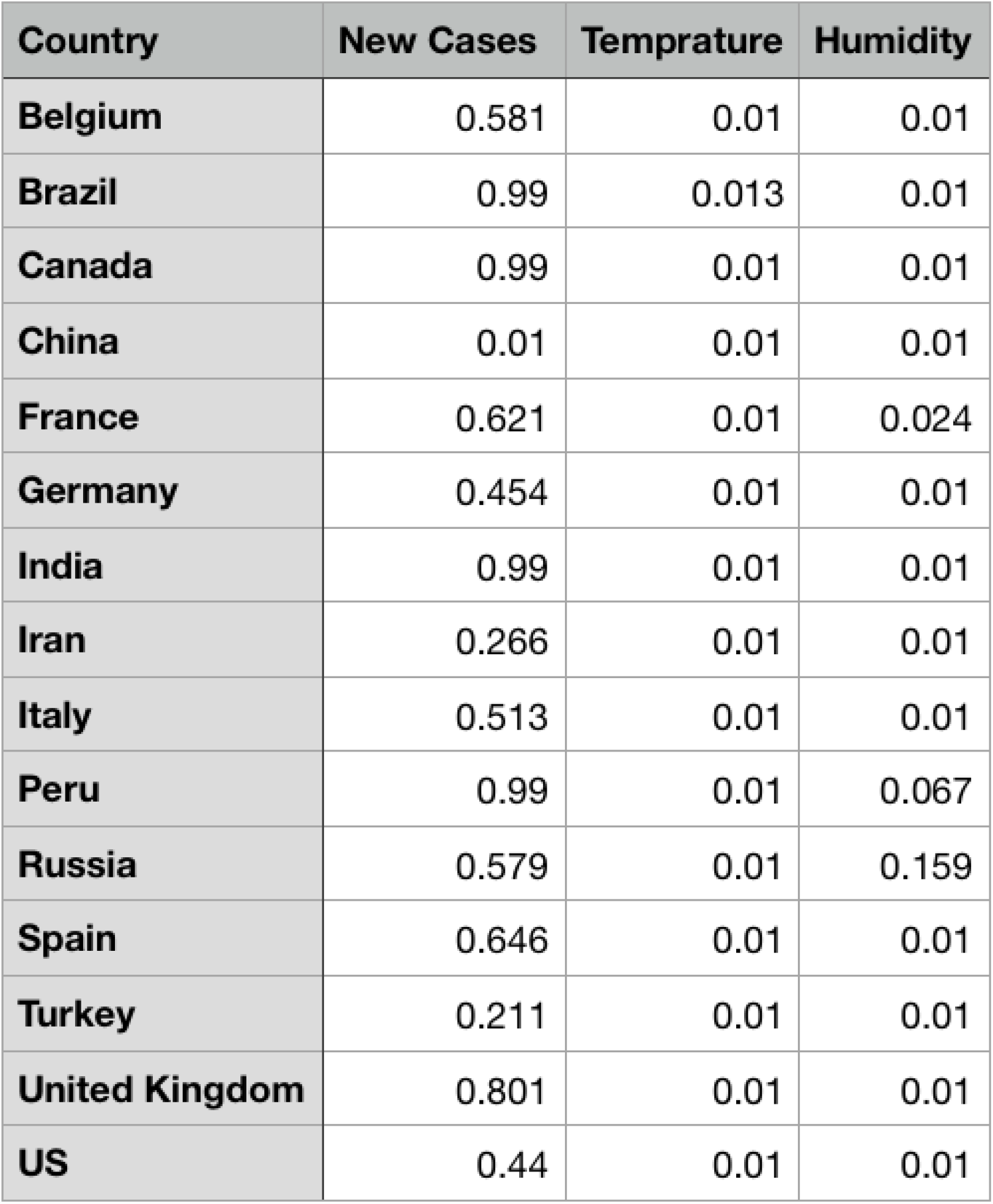
P-value Augmented Dicky Fuller (ADF)Test: Table shows the P-value obtained from ADF test

## References

1. Zhu, Yongjian, and Jingui Xie. “Association between ambient temperature and COVID-19 infection in 122 cities from China.” Science of The Total Environment (2020): 138201.

2. Ma, Yueling, et al. “Effects of temperature variation and humidity on the death of COVID-19 in Wuhan, China.” Science of The Total Environment (2020): 138226.

3. Yao, Ye, et al. “No Association of COVID-19 transmission with temperature or UV radiation in Chinese cities.” European Respiratory Journal (2020).

4. Wang, Jingyuan, et al. “High temperature and high humidity reduce the transmission of COVID-19.” Available at SSRN 3551767 (2020).

5. Bressler, Steven L., and Anil K. Seth. “Wiener–Granger causality: a well established methodology.” Neuroimage 58.2 (2011): 323–329.

6. Penny, William D. “Comparing dynamic causal models using AIC, BIC and free energy.” Neuroimage 59.1 (2012): 319–330.

7. Al-Hussein, Abdul-Basset A., and Fadihl Rahma Tahir. “Epidemiological Characteristics of COVID-19 Ongoing Epidemic in Iraq.”

8. Giordano, Giulia, et al. “Modelling the COVID-19 epidemic and implementation of population-wide interventions in Italy.” Nature Medicine (2020): 1–6.

9. https://science.thewire.in/the-sciences/covid-19-pandemic-infectious-disease-transmission-sir-seir-icmr-indiasim-agent-based-modelling/

10. https://cran.r-project.org/web/packages/RTransferEntropy/vignettes/

11. Noakes, Lyle. “The Takens embedding theorem.” International Journal of Bifurcation and Chaos 1.04 (1991): 867–872.

12. Clark, Adam Thomas, et al. “Spatial convergent cross mapping to detect causal relationships from short time series.” Ecology 96.5 (2015): 1174–1181.

13. Hothorn, Torsten, et al. “Package ‘lmtest’.” (2019).

14. https://cran.r-project.org/src/contrib/Archive/rEDM/

15. https://bmcinfectdis.biomedcentral.com/articles/10.1186/1471-2334-12-298

16. Bressler, Steven L., and Anil K. Seth. “Wiener–Granger causality: a well established methodology.” Neuroimage 58.2 (2011): 323–329.

17. Lin, Jianhua. “Divergence measures based on the Shannon entropy.” IEEE Transactions on Information theory 37.1 (1991): 145–151.

18. Staniek, Matthäus, and Klaus Lehnertz. “Symbolic transfer entropy.” Physical Review Letters 100.15 (2008): 158101.

19. WHO regions: https://www.who.int/healthinfo/global_burden_disease/definition_regions/en/#:~:text=WHO%20regions%3A%20WHO%20Member%20States,Region%2C%20and%20Western%20Pacific%20Region.

